# Transcutaneous Spinal Cord Stimulation to Reduce Phantom Limb Pain in People with a Transtibial Amputation

**DOI:** 10.1101/2023.04.13.23288483

**Authors:** Ashley N Dalrymple, Lee E Fisher, Douglas J Weber

**Author notes:** **Correspondence** Douglas J Weber, 5000 Forbes Ave, Wean 1323 Pittsburgh, PA, USA 15213, Ashley N Dalrymple 20 S 2030 E, BPRB 506D, Salt Lake City, UT, USA 84132.

## Abstract

**Objective:** Phantom limb pain (PLP) is debilitating and affects over 70% of people with lower-limb amputation. Other neuropathic pain conditions correspond with increased spinal excitability, which can be measured using reflexes and F-waves. Spinal cord neuromodulation can be used to reduce neuropathic pain in a variety of conditions and may affect spinal excitability, but has not been extensively used for treating phantom limb pain. Here, we propose using a non-invasive neuromodulation method, transcutaneous spinal cord stimulation (tSCS), to reduce PLP and modulate spinal excitability after transtibial amputation.

**Approach:** We recruited three participants, two males (5- and 9-years post-amputation, traumatic and alcohol-induced neuropathy) and one female (3 months post-amputation, diabetic neuropathy) for this 5-day study. We measured pain using the McGill Pain Questionnaire, visual analog scale, and pain pressure threshold test. We measured spinal reflex and motoneuron excitability using posterior root-muscle (PRM) reflexes and F-waves, respectively. We delivered tSCS for 30 minutes/day for 5 days.

**Main Results:** After 5 days of tSCS, pain scores decreased by clinically-meaningful amounts for all participants from 34.0±7.0 to 18.3±6.8. Two participants had increased pain pressure thresholds across the residual limb (Day 1: 5.4±1.6 lbf; Day 5: 11.4±1.0 lbf). F-waves had normal latencies but small amplitudes. PRM reflexes had high thresholds (59.5±6.1 µC) and low amplitudes, suggesting that in PLP, the spinal cord is hypoexcitable. After 5 days of tSCS, reflex thresholds decreased significantly (38.6±12.2 µC; p<0.001).

**Significance:** Overall, limb amputation and PLP may be associated with reduced spinal excitability and tSCS can increase spinal excitability and reduce PLP.

## INTRODUCTION

Over 3.6 million people will be living with a lower-limb amputation in the United States by the year 2050 (Ziegler-Graham et al., 2008). Following a lower-limb amputation, over 70% of people experience phantom limb pain (PLP) (Ephraim et al., 2005; van der Schans et al., 2002; Ziegler-Graham et al., 2008). Pain in the residual limb and phantom sensations are also common following limb amputation (Hsu and Cohen, 2013; Urits et al., 2019). PLP can be described as sharp, shooting, squeezing, burning, itching, piercing, dull, tingling, throbbing and/or cramping (Hsu and Cohen, 2013; Jensen et al., 1985; Petersen et al., 2019). PLP is menacing and significantly reduces the quality of life of those who suffer from it (van der Schans et al., 2002), disrupting sleep, appetite, ability to focus, hygiene, socialization, and mood (Padovani et al., 2015).

Treatments for PLP include pharmaceuticals, surgical interventions, and mirror therapy. Mirror therapy involves looking at the reflection of voluntary movements made by the intact limb, creating a visual illusion of a non-painful, moving limb in place of the missing limb (Petersen et al., 2019). While there are many reports of the success of mirror therapy (Chan et al., 2007; Finn et al., 2017), a systematic review highlighted a lack of evidence for its efficacy (Barbin et al., 2016). Pharmacologic treatments may include local anesthetics, amitriptyline, duloxetine, acetaminophen, gabapentin, and opioids (Urits et al., 2019). Pharmacologic treatments often lose efficacy over time, have serious side effects, and can lead to addiction (Hsu and Cohen, 2013; Park et al., 2022). Surgical interventions are typically explored when other treatment methods have been deemed ineffective (Davis, 1993). Surgical interventions such as a cordotomy (Perneczky and Sunder-Plassmann, 1975; Pool, 1946), tractotomy (Nikolajsen and Jensen, 2001), dorsal root entry zone lesioning (Saris et al., 1985; Tomycz and Moossy, 2011), and coblation of peripheral nerves (Zeng et al., 2016) are permanent solutions with varied efficacy for PLP (Hu et al., 2007; Tomycz and Moossy, 2011), and can also result in a loss of function, such as a further loss of sensation (Chalil et al., 2021; Ruiz-Juretschke et al., 2011).

Neuromodulatory therapies using electrical stimulation have been viewed as a last resort for treating PLP (Petersen et al., 2019; Urits et al., 2019). Transcutaneous electrical nerve stimulation (TENS) is a non-invasive neuromodulatory technique in which electrical stimulation is delivered through adhesive electrodes placed on the surface of the skin near the pain site and has been shown to relieve PLP, but is more effective for stump pain (Mulvey et al., 2013; Tilak et al., 2016). Epidural spinal cord stimulation (eSCS) of the dorsal columns has been shown to reduce PLP (Aiyer et al., 2017; Krainick et al., 1980, 1975; Viswanathan et al., 2010). It has been suggested that lateral eSCS, targeting the dorsal spinal roots, can provide more effective pain relief (Bunch et al., 2015; Nanivadekar et al., 2022a). Furthermore, there is evidence that dorsal root ganglion (DRG) stimulation can effectively target pain in the distal limbs, including PLP (Eldabe et al., 2015; Esposito et al., 2019).

The dorsal spinal cord and DRG are targets-of-interest because these structures undergo neuroplastic changes with chronic and neuropathic pain. Repetitive activation of nociceptive fibers in the peripheral nerves resulting from the nerve transection induces windup in the dorsal horn neurons, increasing their excitability (Woolf, 1983; Woolf and Thompson, 1991). Furthermore, sprouting of axotomized nerve fibers in the dorsal horn contributes to allodynia and hyperalgesia (Melzack et al., 2001; Woolf et al., 1992). After peripheral nerve injury, DRG neurons ectopically discharge and are hyperexcitable due to changes in ion channel expression and sprouting (McLachlan et al., 1993; Vaso et al., 2014; Wall and Devor, 1983). An increase in spinal cord excitability has also been reported in the absence of sensation (Thompson et al., 2019), following peripheral nerve injury (Valero-Cabré and Navarro, 2001), and in painful diabetic neuropathy (Lee-Kubli et al., 2018; Lee-Kubli and Calcutt, 2014; Marshall et al., 2017). Hoffman (H)-reflexes, evoked by stimulating the tibial nerve and recorded in the soleus muscle, were prolonged or absent in diabetic neuropathy (Millán-Guerrero et al., 2012), but are hyperexcitable in painful diabetic neuropathy (Lee-Kubli et al., 2018; Lee-Kubli and Calcutt, 2014; Marshall et al., 2017). To date, H-reflexes have only been reported in people with limb amputation using implanted cuff electrodes, as opposed to traditional methods that use surface electrodes (Li et al., 2023). A related reflex, the posterior root-muscle (PRM) reflex, is evoked by stimulating the dorsal spinal roots. The PRM reflex is a superposition of the H-reflex and cutaneous afferent inputs (Freitas et al., 2022; Krenn et al., 2013; Minassian et al., 2007), and can also be used to measure spinal excitability (Minassian et al., 2020).

Transcutaneous spinal cord stimulation (tSCS) is a non-invasive neuromodulation method that targets the dorsal spinal roots, similar to eSCS (Dalrymple et al., 2023; Hofstoetter et al., 2018). tSCS can be used to evoke PRM reflexes, and can potentially be used as a neuromodulation therapy. To date, tSCS has been used to improve motor recovery after spinal cord injury (Hofstoetter et al., 2013; Inanici et al., 2021; Keller et al., 2021; Sayenko et al., 2019) as well as to reduce spasticity (Hofstoetter et al., 2020, 2014; Knikou and Murray, 2019). tSCS has not yet been tested as a therapy for chronic or neuropathic pain.

In this study, we elicited reflexes and F-waves to determine if the spinal cord of people with a lower-limb amputation and PLP had altered excitability. We hypothesized that spinal reflexes would be hyperexcitable, indicated by lower thresholds to evoke the reflexes, because of the neuropathic pain state. We also measured F-waves, which indicate motoneuron excitability (Kane and Oware, 2012; Nobrega et al., 2004). We hypothesized that, similar to the reflexes, the motoneurons would be hyperexcitable due to the neuropathic pain state. However, F-wave analysis in neuropathic pain models has not been reported to the best of our knowledge. We applied tSCS each day for 5 days, targeting the dorsal roots corresponding to the distal limbs. We hypothesized that, after 5 days of tSCS, spinal reflex hyperexcitability and PLP would decrease.

## MATERIALS AND METHODS

### Participants

Three individuals with a unilateral transtibial amputation participated in this study (Table 1). We excluded individuals from this study if they were younger than 18 years of age, were pregnant, or had any of the following: implanted electronic devices, any serious disease, disorder, infection, or cognitive impairments, a history of spinal cord injury, herniated disk, or myelopathy, or heart disease including arrhythmia. This study was approved by the Internal Review Board at Carnegie Mellon University and conducted in accordance with the Declaration of Helsinki. All participants provided written informed consent prior to their enrollment in the study. No participants had prior experience with tSCS. One participant (Participant 1) had received 30 days of eSCS more than two years prior, as part of a study using eSCS to restore sensation in the missing limb (Nanivadekar et al., 2022a). All participants had tried and abandoned TENS. The current study took place over 5 days in one week. Participants sat comfortably in a chair or their wheelchair (Participant 2, who was not yet fit for a prosthesis) and removed their prosthesis (Participants 1 and 3). All 3 participants were taking pain medication, including gabapentin (all participants), morphine (Participant 1), hydrocodone (Participant 1), and oxycodone (Participant 3). When they enrolled in the study, they were not asked to alter their medication regimen.

**Table 1.** Demographic information for research participants.

| Participant ID | Age | Gender | Time since amputation | Nature of amputation | Side of amputation |
| --- | --- | --- | --- | --- | --- |
| 1 | 56-60 | M | 9 years | Traumatic | Left |
| 2 | 36-40 | W | 3 months | Diabetic neuropathy | Right |
| 3 | 46-50 | M | 5 years | Alcoholic neuropathy | Left |

### Pain Measures

Participants completed the Groningen Questionnaire Problems after Leg Amputation (GQPLA), which is a qualitative survey intended to gain insight into difficulties that may arise following a leg amputation (van der Schans et al., 2002). The GQPLA is a modified version of a similar questionnaire for people with upper-limb amputations (Kooijman et al., 2000). The GQPLA asks participants to describe their phantom sensations, PLP, and stump pain. It also characterizes changes in prosthesis use (Supplementary Table 1).

We performed the pain pressure threshold (PPT) test using an algometer (Wagner Instruments, Greenwich, CT, USA). The PPT test measures the minimum amount of pressure that the participant can tolerate at a specific location. We pressed the rubber tip (1 cm diameter) of the algometer onto the skin over muscle (not pushing on bone) on several locations of both the residual and intact limbs (Table 2). Participants reported when the pressure became painful, at which point we removed the algometer and recorded the pressure magnitude.

**Table 2.** Locations where the pain pressure threshold test was performed.

| Residual Limb | Intact Limb |
| --- | --- |
| Bottom of stump | Bottom of heel |
| Anterior 5 cm above stump | Ball of foot |
| Posterior 5 cm above stump | Dorsum of foot |
| Medial 5 cm above stump | Posterior ankle |
| Lateral 5 cm above stump | Mid-shin |
| Anterior 10 cm above stump | Mid-calf |
| Posterior 10 cm above stump | Mid-quadriceps |
| Medial 10 cm above stump | Mid-hamstring |
| Lateral 10 cm above stump |  |
| Mid-quadriceps |  |
| Mid-hamstring |  |

We asked participants to rate their PLP in the last 24 hours using a visual analog scale (VAS) from 0 and 10, where 0 indicated no pain at all, and 10 indicated the worst pain imaginable. Participants completed the short-form McGill Pain Questionnaire (MPQ) to describe their pain prior to participation in the study as well as throughout the week. The MPQ evaluates the sensation, temporal changes, and strength of pain. The total MPQ score indicates the intensity and affect the pain has on their life.

### Eliciting Peripheral and Spinal Reflexes

To measure spinal cord excitability, we studied motor (M)-waves, F-waves, and PRM reflexes, which were recorded using electromyography (EMG) electrodes placed on the residual limb. Prior to placing the EMG electrodes, we prepared the skin on the residual limb using abrasive gel (Lemon Prep, Mavidon, USA), alcohol wipes (Braha Industries, USA) and conductive electrode gel (Signa Gel, Parker Laboratories BV, NL). We placed bipolar electromyography (EMG) electrodes (Dual foam Ag|AgCl electrode, 7/8”×1 5/8”, MVAP Medical Supplies, Thousand Oaks, USA) on the lateral gastrocnemius (LG), medial gastrocnemius (MG), tibialis anterior (TA), and vastus lateralis (VL) muscles and a high-density EMG electrode grid (large, 64 channel; TMSi, NL) across the putative gastrocnemius muscles (Figure 1a). The locations of these muscles were confirmed using palpation during attempted movements of the missing ankle. We positioned a ground electrode (4×5 cm pregelled Ag|AgCl Natus electrode; MVAP Medical Supplies, Thousand Oaks, USA) onto the patella of the residual limb. After the electrodes were placed, we wrapped the residual limb with an elastic bandage (Coban, 3M, St. Paul, USA) to reduce swelling that can occur with the prosthesis off. We recorded EMG using the SAGA64+ (TMSi, NL) at a sampling rate of 4000 Hz and streamed the data into MATLAB (MathWorks, USA).

**Figure 1:**
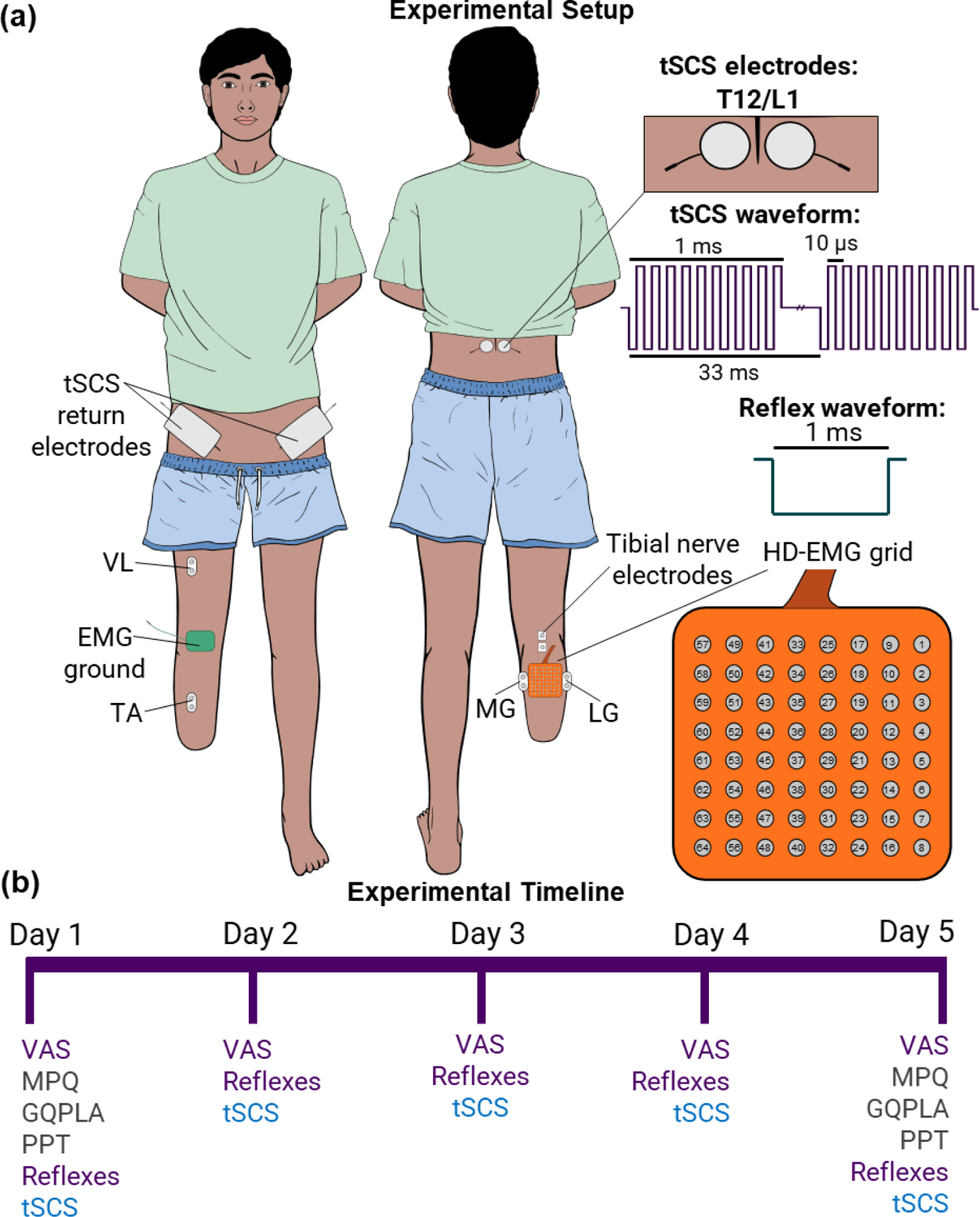
(a) Graphic of participant with a transtibial amputation with electrode placement indicated. Transcutaneous spinal cord stimulation (tSCS) electrodes were placed paraspinally of the T12/L1 vertebrae, return electrodes were placed bilaterally on the anterior superior iliac spines. Bipolar electromyography (EMG) electrodes were placed on the vastus lateralis (VL), tibialis anterior (TA), medial gastrocnemius (MG), and lateral gastrocnemius (LG) muscles. A 64-channel high-density electromyography (HD-EMG) grid was placed on the putative gastrocnemius muscles. High-frequency tSCS waveform and reflex waveform are shown. (b) Study timeline across the 5 days. Intervention and metrics: VAS = visual analog scale; MPQ = McGill Pain Questionnaire; GQPLA = Groningen Questionnaire Problems after Leg Amputation; PPT = Pain Pressure Threshold test; Reflexes refers to both PRM reflexes, M-waves, and F-waves; tSCS refers to 30 minutes of high-frequency tSCS.

We delivered stimulation using a DS8R stimulator with a firmware update to allow frequencies up to 10 kHz (Digitimer, UK). We used custom MATLAB software to control the amplitude and timing of stimulation pulses via analog and digital outputs generated by a multifunction I/O device (PCIe 6353; National Instruments, USA). We elicited M-waves and F-waves by electrically stimulating the tibial nerve of the residual limb. We first confirmed the location of the tibial nerve using ultrasound imaging (Butterfly Network, Inc., Burlington, MA, USA), then placed electrodes (2 square 7/8”×7/8” Ag|AgCl foam electrodes; MVAP Medical Supplies, Thousand Oaks, USA) longitudinally in the popliteal fossa with approximately 1-2 cm spacing. Stimuli consisted of a 1-ms long monophasic, cathodic, square wave pulse. We varied the stimulation amplitude to determine the thresholds for the M-wave as well as the maximum amplitude of the M-wave (M_MAX_). The M_MAX_ was at the stimulation amplitude past which the magnitude of the M-wave no longer increased. We evoked F-waves by stimulating supramaximally (Jerath and Kimura, 2019; Lee et al., 2004).

We elicited the PRM reflex in the residual limb by electrically stimulating the spinal dorsal roots using tSCS (Dalrymple et al., 2023). We placed round adhesive electrodes (3.2 cm diameter; ValuTrode, Axelgaard Manufacturing Co. Ltd., USA) paravertebrally of the T12-L1 spinous processes (Figure 1a). The placement of the tSCS electrodes was chosen to specifically target the dorsal roots corresponding to sensorimotor pathways innervating the distal leg muscles (Calvert et al., 2019; Krenn et al., 2013). We placed return electrodes on bilateral anterior superior iliac spines (7.5×13 cm, ValuTrode, Axelgaard Manufacturing Co. Ltd., USA). We wrapped the participant’s torso using Coban wrap (6”, 3M, USA) and placed a small piece of foam (12×17 cm) between the tSCS electrodes and the back of the chair. These last steps were to ensure firm pressure was maintained on the stimulation site. The stimulation waveform used to evoke the PRM reflex was the same as for the M-wave: a 1-ms long monophasic, cathodic, square wave pulse. For evoking PRM reflexes in the residual limb, we stimulated through the tSCS electrode ipsilateral to the residual limb only. We determined the stimulation threshold for evoking a PRM reflex in the gastrocnemius muscles, followed by the maximum PRM reflex amplitude (PRM_MAX_). The maximum PRM reflex amplitude was at either the stimulation amplitude past which the magnitude of the PRM reflex no longer increased or the maximum stimulation amplitude tolerated by the participant. In this study, we did not exceed a stimulation amplitude of 180 mA.

Our primary muscles-of-interest were the MG and LG. We recorded from the VL and TA muscles to guide tSCS electrode placement to ensure targeting of the distal muscles.

We varied the stimulation amplitude to obtain recruitment curves for the M-waves, F-waves, and PRM reflexes. Specifically, we stimulated at 15 levels between 5 mA below threshold and 10-15 mA above the M_MAX_ (or PRM_MAX_; if tolerated or up to 180 mA) in a random order. Each amplitude was repeated four times and stimuli were delivered 10 s apart.

### Transcutaneous Spinal Cord Stimulation

We delivered bilateral tSCS for neuromodulation continuously for 30 minutes, with a break at 15 minutes to remove the electrodes and inspect the stimulation site. tSCS consisted of a 1 ms-long biphasic burst of pulses at 10 kHz (50 µs/phase; known as a carrier frequency), delivered at 30 Hz (Figure 1a). We started with a low amplitude of stimulation (approximately 10-20 mA) and slowly increased the stimulation beyond PRM reflex threshold, according to the comfort of the participant (maximum tolerated = 90, 110, and 135 mA for Participants 1, 2, and 3, respectively). This ramping up process was completed in less than 2 minutes.

### Study Protocol

All data were collected while the participant sat comfortably in a chair. At the beginning of each day, the participant rated their PLP over the last 24 hours using the VAS. We marked the location of the stimulation and EMG electrodes with a permanent marker to ensure consistent placement across the 5 days. Every day, we performed the M-wave, F-wave, and PRM reflex measures, followed by high-frequency tSCS for 30 minutes. At the beginning of the first and fifth days, the participant completed the MPQ and GQPLA, and we performed the PPT test. The complete study timeline is shown in Figure 1b.

### Analyses and Statistics

We determined the change in PPT over time by subtracting the PPT value at each location on Day 1 from Day 5. We normalized the PPT at each location to the maximum value recorded for each participant. We expressed the change in PPT on a scale between −1 and 1, where −1 indicated the maximum decrease in PPT, 1 indicated the maximum increase in PPT, and 0 indicated no change. We compared the average PPT across all locations on each limb between Days 1 and 5 using a paired t-test. We used repeated measures ANOVA to compare the mean VAS score from all participants across the 5 days. A decrease in the VAS score by 50% and at least 1 point is considered clinically meaningful. We summated the responses from each subsection of the MPQ to obtain a total score. A clinically meaningful decrease in MPQ score is 5 points (Strand et al., 2008).

We removed the stimulus artefact in the EMG signals by interpolating between pre- and post-stimulus time intervals. For analyzing PRM reflexes, the artefact blanking window began 4 ms prior and ended 6 ms after the stimulus onset. The response window was between 10 and 40 ms post-stimulus. The response window for the M- and F-waves was 5 to 60 ms. For analyzing M-waves and F-waves, the blanking interval was between 4 ms prior to and 9 ms after the stimulus onset. The post-stimulus blanking period was larger for the M-wave and F-wave analysis because this interval provided the clearest M-wave. However, the M-wave was contaminated by the stimulus artefact, which was removed by blanking; therefore, M-wave latency could not be accurately measured. All EMG data were filtered using a 2^nd^ order Butterworth bandpass filter with cutoff frequencies at 20 Hz and 1999 Hz.

We defined the threshold for evoking a PRM reflex or M-wave as the lowest stimulation amplitude that elicited a response that was three standard deviations beyond the mean baseline (pre-stimulus) period and confirmed this visually. Grouped threshold data contained the mean threshold across all channels of the HD-EMG grid. We expressed reflex thresholds in units of charge (µC), obtained by multiplying the stimulation amplitude (in mA) by the pulse width (1 ms). Note that when evoking reflexes, the 10 kHz carrier frequency was not applied. We performed a linear correlation between the PRM reflex threshold for each electrode on the HD-EMG grid with its impedance value to determine if any variability in threshold values related to their spatial distribution over the residual limb muscles.

We determined the latency of the F-waves to be the time from the onset of the stimulation to the first inflection of the response. The first inflection was detected when the amplitude of the response exceeded two standard deviations beyond the mean baseline period. We measured the duration of the F-wave, which we defined as the time from the onset of the F-wave to the offset. We defined the offset as the time when the response returned to baseline. To compare the PRM reflex thresholds over 5 days, we performed a one-way analysis of variance (ANOVA) with Bonferroni post-hoc tests.

We created recruitment curves for the PRM reflexes, M-waves, and F-waves by plotting the mean peak-to-peak amplitude of the response from each electrode on the HD-EMG grid as a function of stimulation amplitude. We determined the slope of the recruitment curve, or recruitment rate, for the PRM reflexes using the MATLAB function *findchangepts* to find the inflection points of the mean curve for each participant. We set the parameters for the *findchangepts* function as follows: the maximum number of change points equal to two; the minimum allowable number of samples equal to two. The slope of the recruitment curve corresponded to the slope of the line between the inflection points across the steepest part of the curve. We calculated the F/M ratio by dividing the maximal peak-to-peak amplitude of the F-wave (F_MAX_) by the M_MAX_ and expressed the F/M ratio as a percentage. We compared the change in the F/M ratio, F-wave latency and duration between Day 1 and Day 5 for each Participant using a t-test.

For all comparisons, we tested for normality using the Shapiro–Wilk test and assessed the homogeneity of variance using Levene’s test. A p-value < 0.05 was considered significant.

## RESULTS

Through the GQPLA, all participants reported experiencing recurring leg pain that required medication prior to amputation, ranging from one week to one year in duration. All participants sought previous treatment for PLP and stump pain, including pain medications. Participants 1 and 3 reported some relief with their pain medications, but noted the relief was incomplete. Participant 1 underwent a scar revision surgery and a neuroma surgery and reported temporary relief from those procedures and with medication. Over the course of the study, there were no changes in prosthesis use by Participants 1 and 3 (Participant 2 did not have a prosthesis yet). Both participants reported using their prosthesis daily, for 8 hours or more, and were capable of walking distances of 0.5 miles or longer.

tSCS neuromodulation was well tolerated by all participants, with no adverse events. Inspection of the skin at the stimulation site did not reveal any redness or irritation. Typically, the initial increase in stimulation amplitude was felt by the participant, and after a few seconds, their awareness of the stimulation subsided.

### tSCS Reduced the Frequency of Stump Pain and PLP

We used the GQPLA to gain understanding of the participants’ experiences with phantom sensations, PLP, and stump pain separately. Reponses revealed that Participants 1 and 2 experienced phantom sensations a few times per day both before and during their participation in the study. Participant 3 reported a reduction in phantom sensations from a few times per hour to a few times during the week of the study. All participants reported feeling itching sensations, while individual phantom sensations included warmth, movement, electric sensations, touch, and abnormal position. Stump pain moderately affected all participants. Participant 1 had no change in the frequency of their stump pain throughout the study; however, Participants 2 and 3 reported reduced frequency of stump pain, from a few times per week to less than once per week and a few times per day to not at all, respectively. All participants reported a reduction in episodes of PLP. Participants 1 and 2 reported experiencing PLP a few times per day before the study, but only a few episodes during the week of the study. Participant 3 reported experiencing PLP a few times per hour before the study, and similarly only experiencing PLP a few times during the week of the study.

### tSCS Reduced Pain Scores

VAS scores for each participant, reflecting phantom limb pain experienced over the previous 24 hours, varied throughout the 5 days (Figure 2a). Participant 1 had an initial score of 6, which decreased steadily to 4.5 by Day 5; however, this difference did not meet the clinically meaningful threshold. Participant 2 had an initial and final score of 4, but on Day 3 reported a score of 0, which is clinically meaningful, but a final score of 4. Participant 3 had a score of 8 on the first day, which decreased to 5 on the final day. Notably, on Day 4, they reported a score of 4, which met the clinically meaningful threshold. The reduction in mean VAS scores across the 5 days was not clinically meaningful and nor statistically significant (p = 0.56).

**Figure 2.**
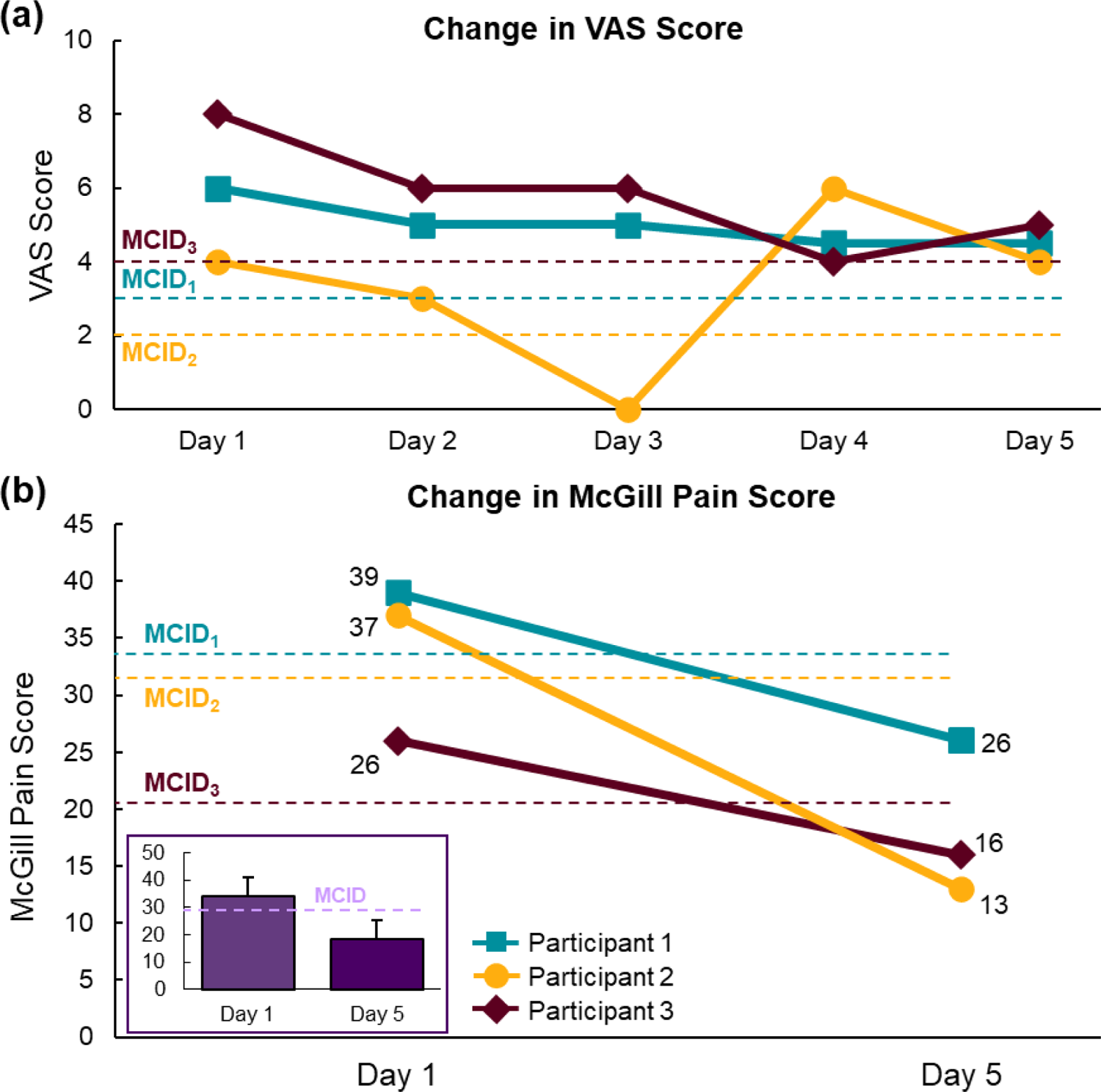
Change in pain scores across the 5 days of using transcutaneous spinal cord stimulation for each participant. (a) Visual analog scale (VAS) score across 5 Days. (b) Scores from the McGill Pain Questionnaire on Days 1 and 5. Inset: Mean (+ SD) scores for all participants. MCID = minimal clinically-important difference.

All participants had clinically meaningful different MPQ scores between Days 1 and 5. The MPQ for Participant 1 decreased from 39 to 26; Participant 2 decreased from 26 to 16; and Participant 3 decreased from 37 to 13 (Figure 2b). As a group, mean MPQ scores decreased from 34.0 (± 7.0) on Day 1 to 18.3 (± 6.8) on Day 5.

### tSCS Increased Pain Pressure Threshold

Across the five days of tSCS, two participants had increased pain pressure thresholds across several locations of their residual and intact limbs (Figure 3a). Both participants had significant increases in their pain pressure thresholds on their residual limb (Participant 1: Day 1: 3.8 ± 2.6 lbf, Day 5: 10.4 ± 3.5 lbf, p < 0.001; Participant 2: Day 1: 7.0 ± 2.4 lbf, Day 5: 12.4 ± 7.6 lbf, p = 0.018) (Figure 3b). This means they could tolerate more pressure applied to their residual limb after 5 days of tSCS. Participant 1 had a significant increase in pain pressure threshold on their intact limb (Day 1: 14.9 ± 3.4 lbf, Day 5: 19.1 ± 4.7 lbf, p = 0.003); however, Participant 2 did not (Day 1: 14.0 ± 5.7 lbf, Day 5: 15.0 ± 6.3 lbf, p = 0.62). Participant 3 exhibited a decrease in pain pressure threshold from Day 1 (20.9 ± 4.2 lbf) to Day 5 (13.0 ± 4.0; p < 0.001). This differing result in Participant 3 may be due to his inconsistent use and timing of pain medications on the testing days. On Day 1, he took pain medications 1 hour before the start of testing. On Day 5, he took pain medications 8 hours before the start of testing.

**Figure 3.**
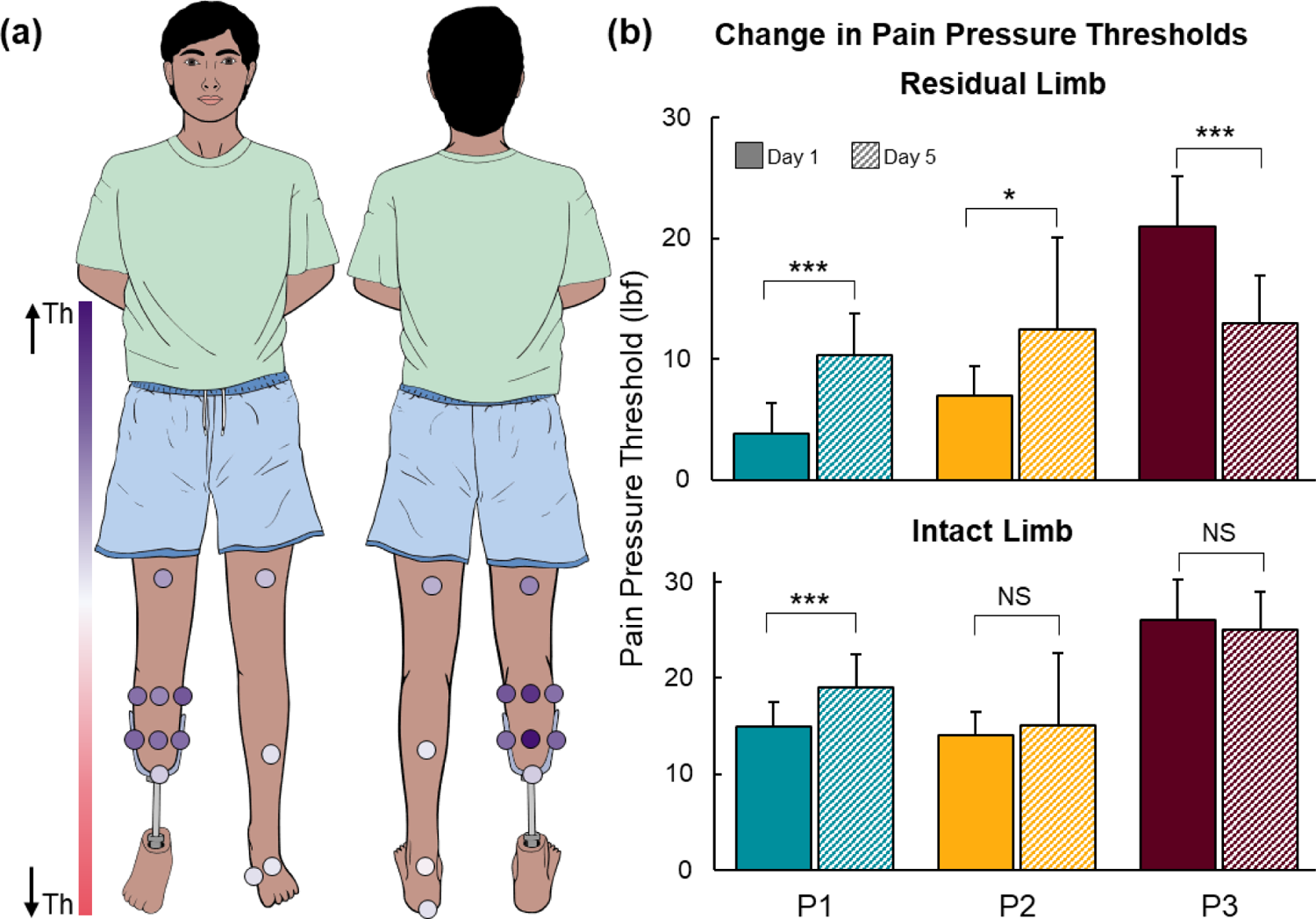
Change in pain pressure threshold (PPT) between Days 1 and 5 of using transcutaneous spinal cord stimulation for Participants 1 (P1) and 2 (P2). (a) Normalized changes in PPT at each location tested on the residual and intact limbs. (b) Mean (+ standard deviation) PPT from all sights tested on the residual limb (top) and intact limb (bottom) on Day 1 (solid) and Day 5 (diagonal lines). Th = threshold; NS = not significant; *p < 0.05; ***p < 0.001.

### Recruitment of PRM Reflexes, M-Waves, and F-Waves Prior to tSCS

Small amplitude PRM reflexes were elicited in the VL and TA muscles in each participant (Supplementary Figure 1). However, the LG and MG muscles were the primary muscles-of-interest because they are the most distal muscles on the residual limb; therefore, our analyses focus on the responses evoked in the putative gastrocnemius muscles. H-reflexes could not be elicited by stimulating the tibial nerve, limiting our analysis to M-waves and F-waves.

The mean PRM reflex thresholds from the HD-EMG grid on the gastrocnemius muscles prior to tSCS neuromodulation were 55.8 ± 8.0 µC, 63.4 ± 1.7 µC, and 59.3 ± 3.7 µC for Participants 1, 2, and 3, respectively (Figure 4; group mean = 59.5 ± 6.1 µC). There was some variability in the PRM reflex thresholds across the electrodes on the HD-EMG grid for Participant 1 (Supplementary Figure 2); however, the thresholds did not correlate with the electrode impedance (r < 0.42).

**Figure 4.**
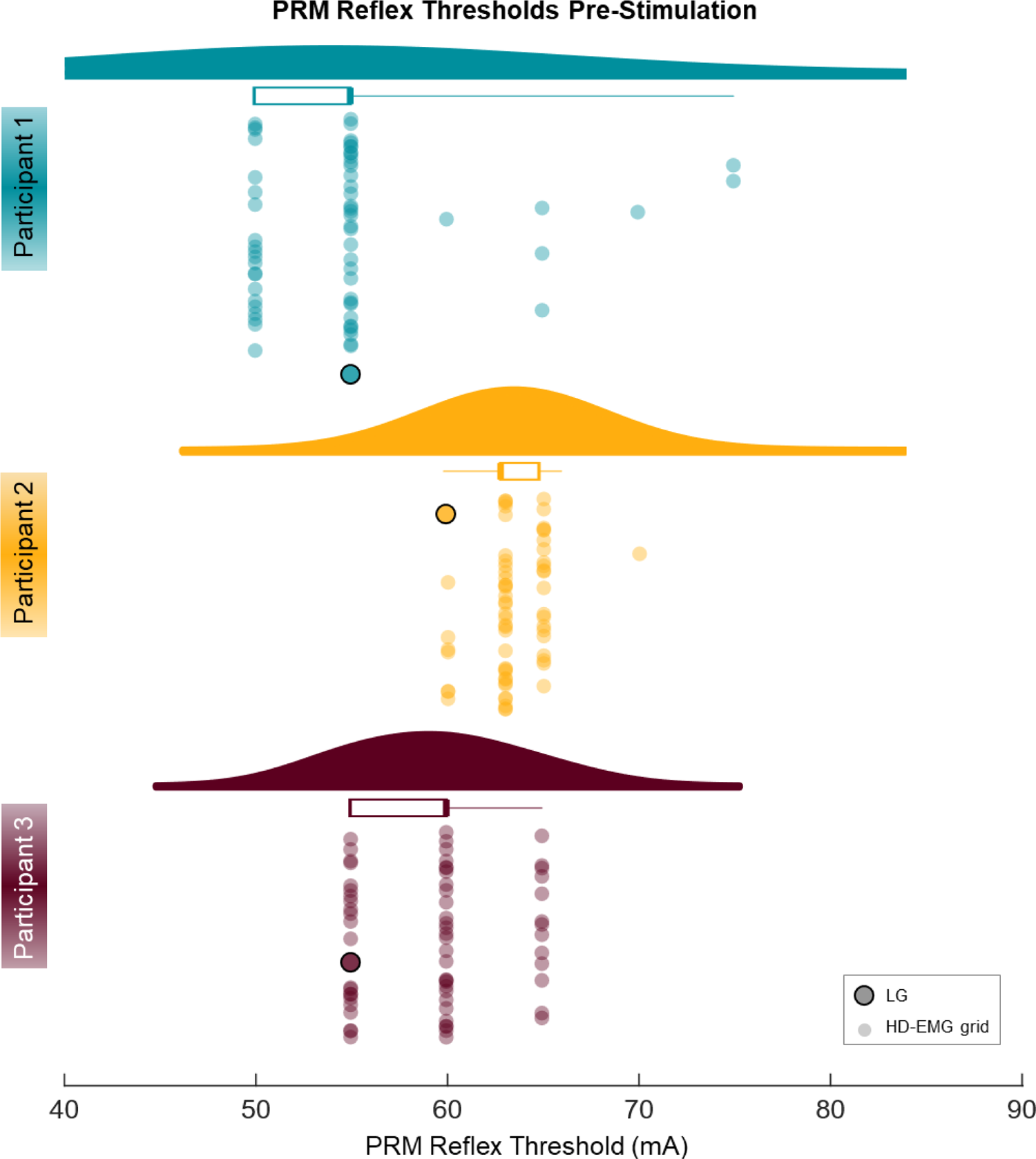
Distribution of stimulation amplitudes required to evoke a posterior root-muscle (PRM) reflex at each electrode on the high-density electromyography (HD-EMG) grid on the putative gastrocnemius muscles and a bipolar EMG recording from the lateral gastrocnemius (LG) muscle. Thresholds were obtained for each participant prior to the use of transcutaneous spinal cord stimulation. The probability density function shows the distribution of the threshold data. The box plot shows the interquartile range and mean of the thresholds. The individual data points were randomly dispersed along the y-axis below the probability density function and box plot, corresponding to the threshold value. The smaller points represent the threshold for each electrode on the HD-EMG grid, and the larger points represent the threshold from the bipolar EMG recording on the LG muscle.

Examples of the PRM reflexes resulting from varied stimulation amplitudes are shown in Figure 5a. The rates of recruitment of the PRM reflexes were 0.028, 0.04, 0.235 mV/µC for Participants 1, 2, and 3, respectively (Figure 5c; group mean = 0.101 ± 0.12 mV/µC). At 2 times the PRM reflex threshold, the peak-to-peak amplitudes of the PRM reflexes were 0.06 ± 0.09 mV (Figure 5d). The recruitment curves are shown in Supplementary Figure 3a. PRM reflexes at 2.5 times threshold for each electrode on the HD-EMG grid for Participant 1 are shown in Supplementary Figure 4.

**Figure 5.**
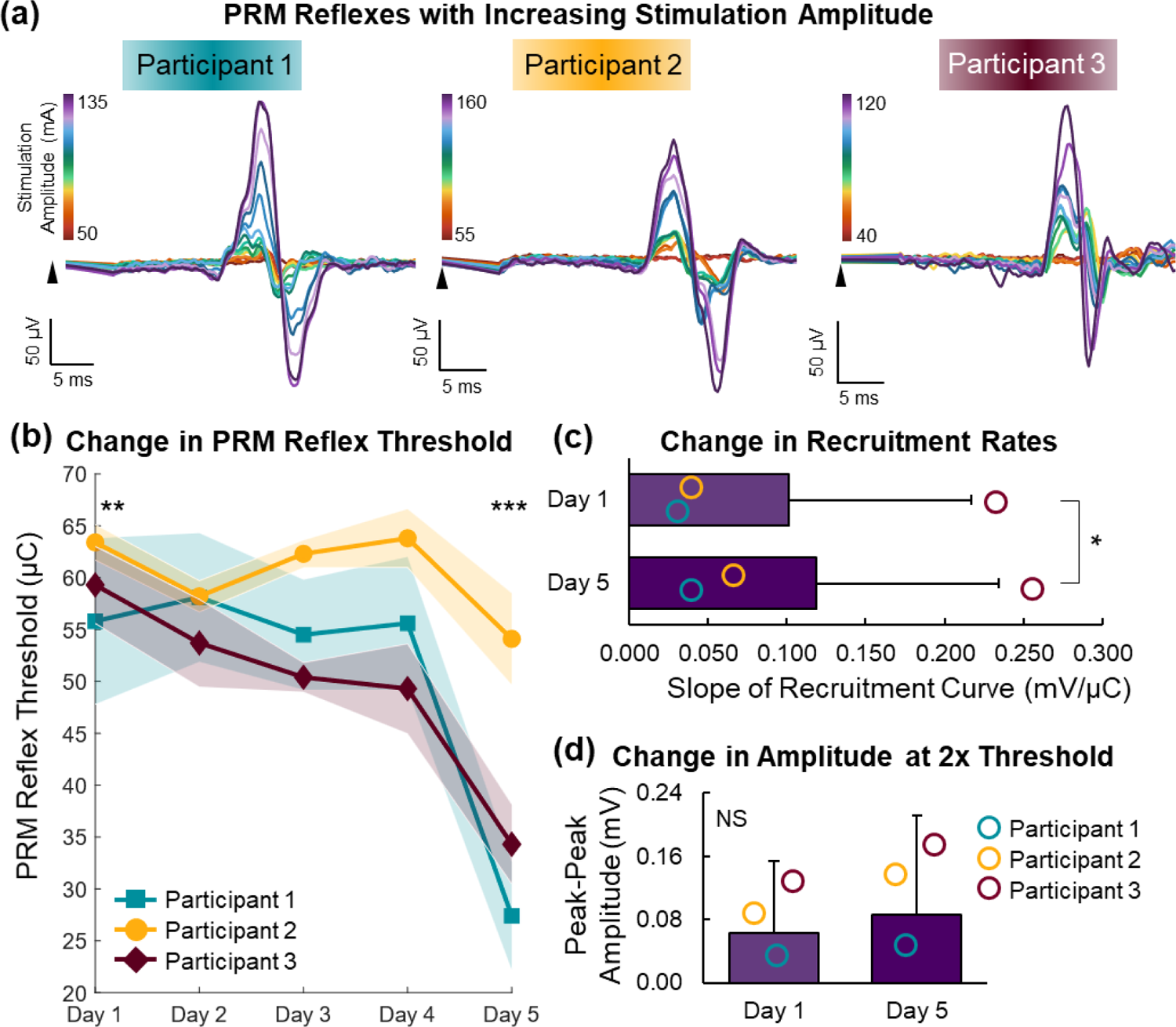
Recruitment of posterior root-muscle (PRM) reflexes. (a) Examples of PRM reflexes from an electrode on the high-density electromyography (HD-EMG) grid as stimulation amplitude was increased. (b) Mean (± standard deviation (SD)) change in amount of charge required to evoke a PRM reflex (threshold). (c) Mean (+SD) slope of the recruitment curves on Days 1 and 5. (d) Mean (+ SD) peak-to-peak amplitude of the PRM reflexes when the stimulation amplitude was 2 times threshold on Days 1 and 5. **p < 0.01; ***p < 0.001.

Examples of the M-waves and F-waves as stimulation amplitude increased are shown in Figure 6a. The recruitment curves from peripheral nerve stimulation show the expected sigmoid of the M-waves (Supplementary Figure 3b). The latencies of the F-waves for Participants 1, 2, and 3 were 38.1 ± 1.5 ms, 44.8 ± 4.1 ms, and 44.4 ± 1.7 ms, respectively (Figure 6c). The latencies for Participants 2 and 3 were within normal range (40 – 64 ms; Jerath and Kimura, 2019; Nobrega et al., 2004). The duration of the F-waves for Participants 1 and 3 were 14.0 ± 3.2 ms and 11.7 ± 2.2 ms, respectively, which are also within the normal range of 13.0 ± 4.5 ms (Nobrega et al., 2004). However, the duration of the F-wave for Participant 2 was 4.8 ± 1.6 ms, which is shorter than normal. F-waves were smaller in amplitude in all participants than those for able-bodied participants (Pereira et al., 2022, 2020). Specifically, the F/M ratios were 4.27 ± 3.10%, 1.19 ± 0.45%, and 4.28 ± 1.02% for Participants 1, 2, and 3, respectively (Figure 6b). This means that on average, the peak-to-peak amplitude of the F_MAX_ was less than 5% of the M_MAX_.

**Figure 6.**
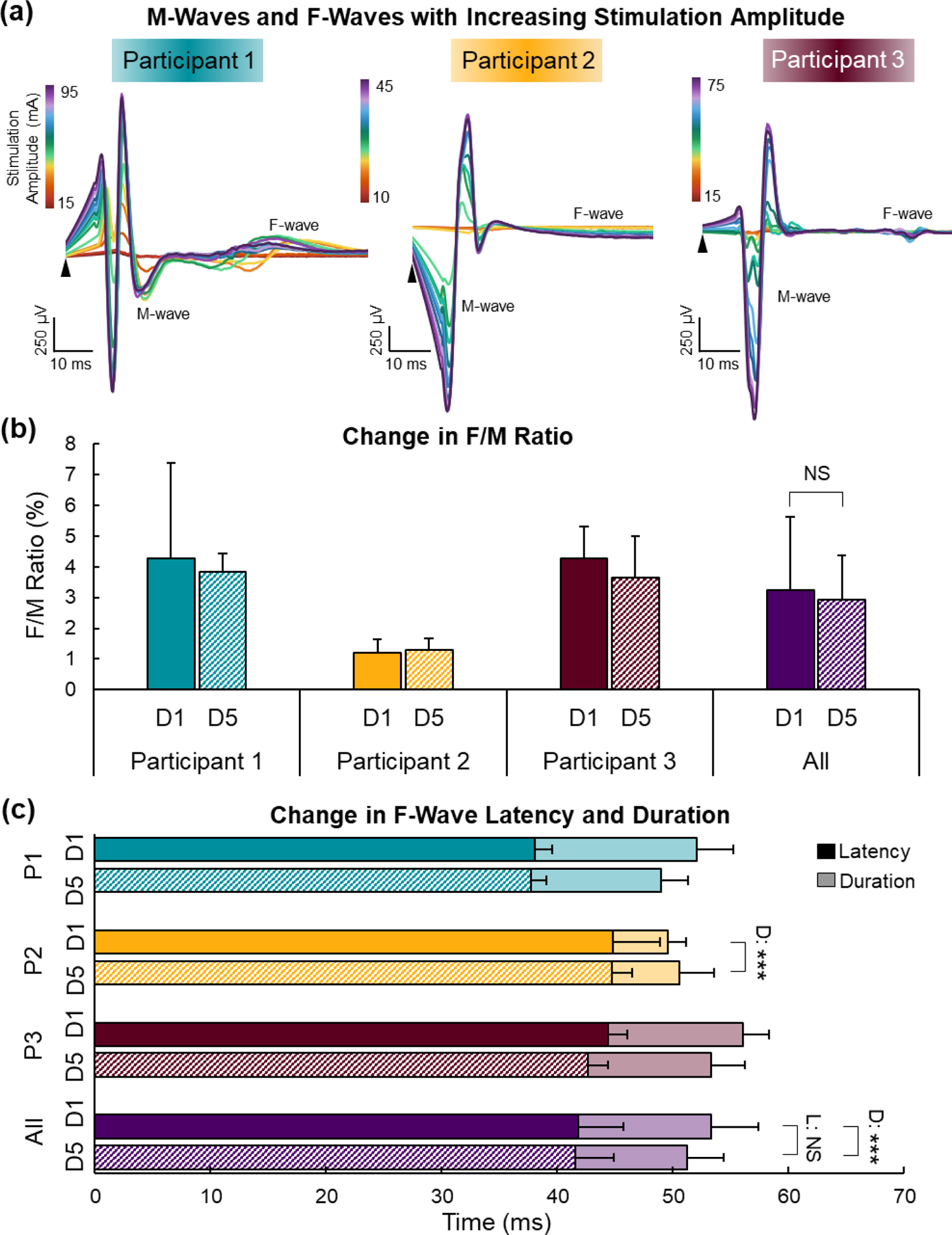
M-waves and F-waves. (a) Examples of M-waves (first response) and F-waves (second, smaller response) from an electrode on the high-density electromyography (HD-EMG) grid as stimulation amplitude was increased. (b) Mean (± standard deviation (SD)) change in ratio of F-wave and M-wave peak-to-peak amplitudes (F/M ratio) across days (D), (c) F-wave latency (L; solid and diagonally striped) and duration (D; semi-transparent) across days for each participant (P). NS = not significant; ***p < 0.001.

### tSCS Decreased PRM Thresholds

After 5 days of tSCS, PRM reflex thresholds decreased in all participants (Figure 5b). Specifically, thresholds for Participant 1 decreased from 55.8 ± 8.0 µC to 27.4 ± 5.2 µC, Participant 2 decreased from 63.4 ± 1.7 µC to 54.1 ± 4.4 µC, and Participant 3 decreased from 59.3 ± 3.7 µC to 34.3 ± 3.8 µC (group means: 59.5 ± 6.1 to 38.6 ± 12.2 µC). The PRM reflex thresholds on Day 1 were significantly higher compared to all other testing days (p < 0.005), and PRM reflex thresholds on Day 5 were significantly lower than on all other days (p < 0.001). The recruitment rates increased significantly after 5 days of tSCS (Day 1: 0.101 ± 0.12; Day 5: 0.119 ± 0.12; p = 0.048; Supplementary Figure 3c; Figure 5c). The peak-to-peak amplitude of the PRM reflexes at 2 times threshold increased over the 5 days, but this increase was not significant (Day 1: 0.06 ± 0.09 mV; Day 5: 0.09 ± 0.13 mV; p = 0.10; Figure 5d).

The F/M ratio did not change significantly between Days 1 and 5 for any participants (Day 1: 3.2 ± 2.4; Day 5: 2.9 ± 1.5; p = 0.11; Figure 6b). Similarly, there were no significant changes in the latency of the F-wave across the 5-days for any participants (Day 1: 41.8 ± 3.9 ms; Day 5: 41.5 ± 3.3 ms; p = 0.44; Figure 6c). However, the duration of the F-waves decreased significantly between Days 1 and 5 but were still within normal range (Day 1: 11.5 ± 4.1 ms; Day 5: 9.7 ± 3.1 ms; p < 0.001). Notably, very few electrodes on the HD-EMG grid detected an F-wave in Participant 2 on Day 1 (26/64 electrodes). However, on Day 5, F-waves were detectable in 55 out of the 64 electrodes (Supplementary Figure 5). Furthermore, although the group data reflected a decrease in F-wave duration over the 5 days, there was a significant increase in F-wave duration in Participant 2, from below normal range to within normal range (p < 0.001; Figure 6c).

### tSCS Evoked Sensory Percepts After a Recent Limb Amputation

Participant 2 had a leg amputation 3 months prior to the study and was not yet fitted for a prosthesis. During the continuous bilateral tSCS for neuromodulation on the first day, and every day thereafter, she reported feeling her missing foot touching the ground. In her own words, “My leg, the amputated one, feels like it’s on the ground, it feels like it’s comfortable. It feels like I have both feet touching the floor. It doesn’t even feel like my foot’s gone.” The sensory percepts were present only during continuous tSCS, not during the brief pulses used for reflex testing. The Participant did not report feeling paraesthesias or other sensations in her intact limb.

## DISCUSSION

Overall, we show that, contrary to our hypotheses, spinal reflex and motoneuron excitability, indicated by PRM reflex recruitment and F-wave characteristics, respectively, are reduced in people who experience PLP. After 5 days of tSCS, spinal reflex thresholds decreased towards those of neurologically intact individuals, and F-waves became more detectable and longer in duration (i.e., closer to normal) in one participant who had a sub-acute amputation. Importantly, 5 days of tSCS reduced the frequency and intensity of stump pain and PLP in all participants. Unexpectedly, tSCS elicited sensory percepts in the missing foot of the participant with a sub-acute amputation.

### Reflex Hypoexcitability with Phantom Limb Pain

Changes in spinal excitability have been characterized following peripheral nerve injury (Valero-Cabré and Navarro, 2001) and for people with diabetes (Lee-Kubli et al., 2018; Millán-Guerrero et al., 2012), but have not been characterized following limb amputation. Two of our participants had neuropathy (diabetic or alcohol-induced) prior to their amputation. In both diabetic neuropathy and alcohol-induced neuropathy, the sensory nerves are affected either by metabolic stress (diabetes) (Feldman et al., 2017) or direct damage (alcohol) (Mellion et al., 2011). We were unable to evoke H-reflexes in any of our participants, regardless of the presence of neuropathy. However, we were able to characterize spinal excitability using PRM reflexes and F-waves, which conduct along the motor nerves in the periphery. Prior to tSCS neuromodulation, PRM reflex thresholds were higher in our participants with transtibial amputations (59.5 ± 6.1 µC) than what we reported in neurologically intact individuals in another study (32.4 ± 9.2 µC; n = 12) (Dalrymple et al., 2023). The initial amplitudes of the PRM reflexes at 2 times threshold were 0.06 ± 0.09 mV, which is more than 130 times smaller than in intact individuals (> 8 mV). The rate of recruitment of PRM reflexes in people with limb amputations (0.101 ± 0.12 mV/µC) was, again, substantially smaller than in intact individuals (∼0.4 mV/µC). The F-waves were extremely small in amplitude, and in the case of Participant 2, who had diabetic neuropathy, were often absent or nearly imperceptible. The amplitudes of the F-waves in all participants were smaller than what has been reported in neurologically intact individuals (Nobrega et al., 2004; Pereira et al., 2022, 2020). The latencies of the F-waves were all within normal range and did not change over time (Jerath and Kimura, 2019). The F-wave duration for Participant 2 was shorter than normal (Nobrega et al., 2004) but increased after 5 days of tSCS. F-wave amplitude and latency can indicate health and excitability of the motoneurons (Kane and Oware, 2012; Nobrega et al., 2004). Collectively, our results indicate that the motoneurons in people with a transtibial amputation may have reduced excitability, evident by the low amplitudes in all participants and small duration in Participant 2.

We characterized PRM reflexes in people with limb amputation and PLP, with and without neuropathy. PRM reflexes were easily obtainable, unlike H-reflexes, in all participants. Therefore, PRM reflexes are a tool that can be used to investigate spinal excitability in populations where the sensory peripheral nerves may be inaccessible or damaged. Overall, prior to tSCS neuromodulation, PRM reflexes had high thresholds, low amplitudes, and low rates of recruitment, suggesting that these spinal reflexes were hypoexcitable. Despite being a neuropathic pain condition, the presence of PLP did not result in spinal hyperexcitability. This indicates that perhaps the limb amputation itself (or perhaps a lack of afferent input) caused spinal hypoexcitability that exceeds any effects from PLP, or that PLP is unique from other chronic and neuropathic pain conditions.

### Plasticity in Sensorimotor Pathways Following Limb Amputation

Following peripheral nerve injury or limb amputation, the primary sensory and motor cortices undergo remapping, where the affected regions have persistent but suppressed cortical representation (Makin et al., 2020; Wesselink et al., 2019). The extent of this cortical remapping correlates with the intensity of PLP, where more severe PLP results in more extensive remapping (Gunduz et al., 2020). Several mechanisms have been proposed to explain cortical remapping, including axonal sprouting in the primary somatosensory cortex (Florence et al., 1998), thalamus, and brainstem (Jones and Pons, 1998), the expansion of the receptive field in the thalamus (Davis et al., 1998; Jain et al., 2008), unmasking or disinhibition of inhibitory connections between somatotopic regions (Li et al., 2014), or unmasking of overlapping receptive fields in the primary somatosensory cortex (Wesselink et al., 2022). Evidence to support the preservation, but suppression, of the canonical topographic cortical map following a limb amputation stems from studies where the nervous system was stimulated and sensory percepts were evoked (Makin et al., 2020). Sensory percepts have been evoked in the missing limbs of people with amputations using electrical stimulation of the peripheral nerves, (Anani and Körner, 1979; Charkhkar et al., 2018; Clippinger et al., 1982; D’Anna et al., 2017; Raspopovic et al., 2014), spinal cord (Chandrasekaran et al., 2020; Nanivadekar et al., 2022a, 2022b), and thalamus (Davis et al., 1998), as well as magnetic stimulation of the contralateral primary motor cortex (Bestmann et al., 2006; Reilly and Sirigu, 2008). Based on these prior studies and the results from our study, indicating spinal hypoexcitability, it is likely that a loss of somatosensory input from an amputation results in a suppression of somatosensory pathways corresponding to the affected limb, and that stimulation ‘reawakens’ or unmasks the sensorimotor nervous system.

### Stimulating the Spinal Cord to Evoke Sensory Percepts

We have previously shown that eSCS can evoke sensory percepts in the missing hand (Chandrasekaran et al., 2020; Nanivadekar et al., 2022b) and foot (Nanivadekar et al., 2022a) following upper- and lower-limb amputation, respectively. eSCS targeting the missing hand resulted in sensory percepts immediately (Chandrasekaran et al., 2020); however, sensory percepts from eSCS targeting the missing foot were absent until approximately two weeks into the stimulation regime (Nanivadekar et al., 2022a). All participants from both studies had chronic (> 2 years) amputations. In the current study, Participant 2, whose amputation was subacute (3 months), reported experiencing sensory percepts in her missing foot immediately following the onset of tSCS on the first day of testing. Participants 1 and 3, whose amputations were chronic, did not report any sensory percepts or paraesthesias during tSCS. It is possible that if we were to deliver tSCS for more than 2 weeks, sensory percepts could be elicited, similar to eSCS. We may have been able to elicit sensory percepts using tSCS following a sub-acute amputation because, even though the spinal cord was hypoexcitable, the somatosensory pathways have not been depressed as extensively as in chronic amputations. Future studies will explore the potential of tSCS as a sensory neuroprosthesis to elicit percepts in the missing limb following acute, sub-acute, and chronic amputations, and will fully characterize the quality of those sensory percepts.

### Clinical Utility and Comparison to Other Methods

As previously mentioned, neuromodulatory therapies that use electrical stimulation are often a last resort for treating PLP (Urits et al., 2019). TENS is better suited for treating stump pain than PLP (Mulvey et al., 2013; Tilak et al., 2016), and eSCS and DRGS require a surgical implant. Here, we proposed a non-invasive method of modulating spinal networks to reduce PLP and stump pain. tSCS is a therapy that could be a more accessible intervention to those who cannot or do not want to undergo a surgical procedure. tSCS could also be used before eSCS or DRGS to determine if the patient will respond to a therapy that targets the dorsal roots. TENS, eSCS, and DRGS are thought to provide pain relief via the gate control theory of pain (Fisher and Lempka, 2023; Graham et al., 2019; Sivanesan et al., 2019), in which the activation of large-diameter Aβ fibers inhibit activity in nociceptive C-fibers in the dorsal horn of the spinal cord (Gibson et al., 2017; Melzack and Wall, 1965). It is possible that tSCS also provides pain relief by activating similar pathways.

tSCS is easy to use, requiring a small number of commercially-available components. Anecdotes from our participants conveyed that the stimulation was tolerable and even unnoticeable after a few minutes. Furthermore, they expressed that they would be willing to use an at-home version of the system as long as the electrode placement could be streamlined. While the stimulation was well-tolerated by our participants, who all had intact sensation on their lower back, it is important to be mindful that electrical stimulation can be uncomfortable for some, and that proper adhesion of the electrodes is important for maintaining comfort. It is also important to consider that contractions of the paraspinal muscles can cause discomfort and a more midline electrode placement can reduce this discomfort (Dalrymple et al., 2023; Manson et al., 2020). We employed a high-frequency carrier of 10 kHz in our stimulation waveform because it has been proposed to be more comfortable than conventional 1 ms-long biphasic stimulation trains (Gad et al., 2017; Gerasimenko et al., 2015; Inanici et al., 2021; Sayenko et al., 2019). However, we recently reported that the addition of a high-frequency carrier does not make tSCS more comfortable, and actually excites spinal reflex pathways less efficiently (Dalrymple et al., 2023).

Many prior studies testing tSCS to restore motor function and reduce spasticity used a conventional waveform (Gad et al., 2017; Hofstoetter et al., 2021; Knikou and Murray, 2019). Both waveforms target the same reflex pathway (Dalrymple et al., 2023), but future studies should be performed to ensure comfort during continuous stimulation with the conventional waveform in people with intact sensation.

### Study Limitations

Although our study included only three participants, we provide preliminary evidence that tSCS may be an effective therapy for reducing PLP in people with a transtibial amputation. The changes in spinal reflex excitability and pain scores occurred over five days, despite the participants’ use of pain medications including gabapentin, opioids, and opiates. Gabapentin is a GABA analogue, which suppresses the nervous system (Rose and Kam, 2002) and may also suppress spinal reflexes, but this has yet to be elucidated. Morphine is an opiate that depresses the central nervous system. It can suppress the monosynaptic spinal reflex but at high concentrations (Mazo et al., 2015). Hydrocodone and oxycodone are opioids that suppress the nervous system, including the nociceptive withdrawal reflex; however, their effects on the monosynaptic reflex is unknown (Fischer et al., 2017). Furthermore, our results show significant and clinically meaningful changes in participants’ pain pressure threshold and MPQ score. A longer duration study may be able to capture a larger change in these measures, as well as a meaningful decrease in VAS score. Additionally, there may be a retention of pain relief provided by tSCS that was not captured in the current study. Future studies will include a follow-up examination of spinal excitability and pain measures to quantify any lasting changes after the cessation of tSCS.

The participants in this study were heterogeneous; they differed in the nature of their amputation, presence of neuropathy, as well as time since amputation. Overall, we demonstrate that people with PLP have spinal hypoexcitability, and that tSCS can increase their spinal excitability and reduce their PLP and stump pain. However, there are individual differences in the extent of altered spinal excitability and changes in reflex recruitment following tSCS. Future work will further investigate the changes in spinal excitability and pain measures following tSCS in a larger cohort, as well as how these changes differ in people with or without a neuropathy, as well as following acute, sub-acute, and chronic amputation. By explicitly investigating the effects of neuropathy and time post-amputation, the commencement and duration of treatment with tSCS may be optimized for maximal pain relief.

The current study did not include a cohort receiving sham stimulation. Any study investigating a treatment for pain must take the placebo effect into consideration. The placebo effect in this case refers to participants reporting pain relief because they know that the therapy is designed to relieve pain, and could confound any true pain relieving effects of the therapy (Colloca, 2019). Pain treatment studies are heavily influenced by participants experiencing the placebo effect (Finsen et al., 1988; Kjær et al., 2020). To account for this, we chose outcome measures that were both subjective and objective. It is possible that the VAS and MPQ scores were influenced by the placebo effect because they rely on the participant subjectively rating their pain throughout the study. However, the changes in PPT and reflex thresholds and recruitment were objective measures, demonstrating that tSCS modulates spinal sensorimotor pathways and reduces hyperalgesia. Nonetheless, future studies should include a group receiving sham stimulation, which can be accomplished by turning stimulation on and slowly reducing the stimulation amplitude to sub-perceptual levels (Rakel et al., 2010).

## CONCLUSIONS

Results from this study suggest that the spinal cord is hypoexcitable in people with transtibial amputations who suffer from PLP, which differs from other chronic and neuropathic pain syndromes. Five days of tSCS reduced the frequency and intensity of stump pain and PLP. Furthermore, tSCS modulated spinal reflex pathways and increased their excitability towards that of neurologically intact individuals. Surprisingly, tSCS evoked sensory percepts in the missing limb of a participant with a sub-acute amputation, suggesting that tSCS could be used as part of a sensory neuroprosthesis. Overall, tSCS is a non-invasive and non-pharmacological neuromodulation method, offering a new hope for people with limb amputations suffering from PLP.

## Data Availability

All data produced in the present study are available upon reasonable request to the authors.

## ACKNOWLEDGMENTS

We greatly appreciate the National Center of National Center for Neuromodulation (NC NM4R) at the Medical University of South Carolina (MUSC) for providing pilot funding for this study. We would like to thank Debbie Harrington, Casey Konopisos, and Alayna Schwerer for their assistance with the IRB protocol, clinical trial registration, and participant recruitment. We are grateful for the advice provided by Dr. Bailey Petersen regarding the placements of the tSCS and EMG electrodes. We would also like to thank Axelgaard for providing us with a suite of electrodes. Finally, we would like to thank our phenomenal research participants for their commitment to science and willingness to participate in this study.

## CLINICAL TRIAL INFORMATION

The funding for this study was registered under clinical trial NCT04543786.

## SUPPLEMENTARY FIGURES

**Supplementary Figure 1.**
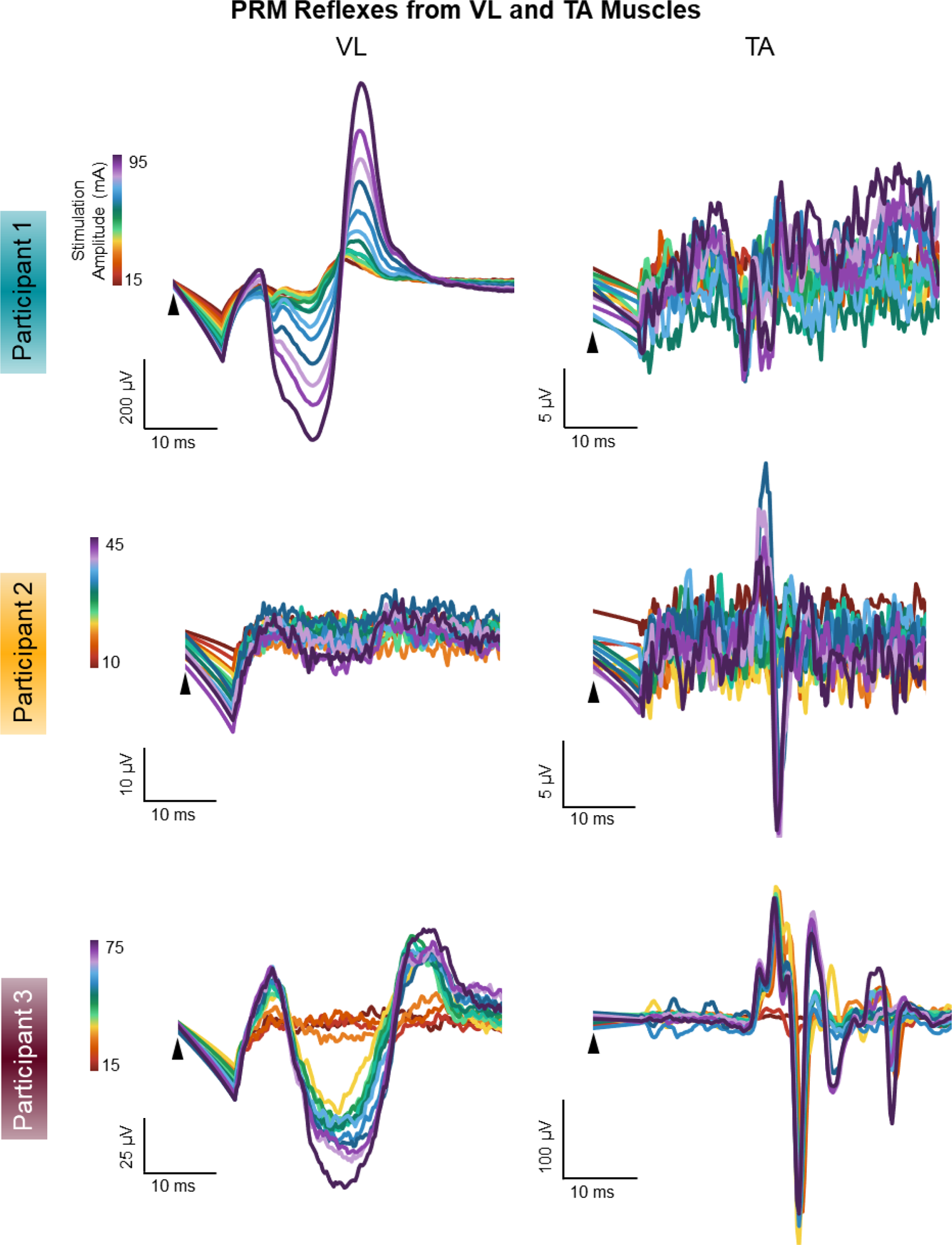
Posterior root-muscle (PRM) reflexes recorded from the vastus lateralis (VL; left) and tibialis anterior (TA; right) muscles of each participant as stimulation amplitude was increased.

**Supplementary Figure 2.**
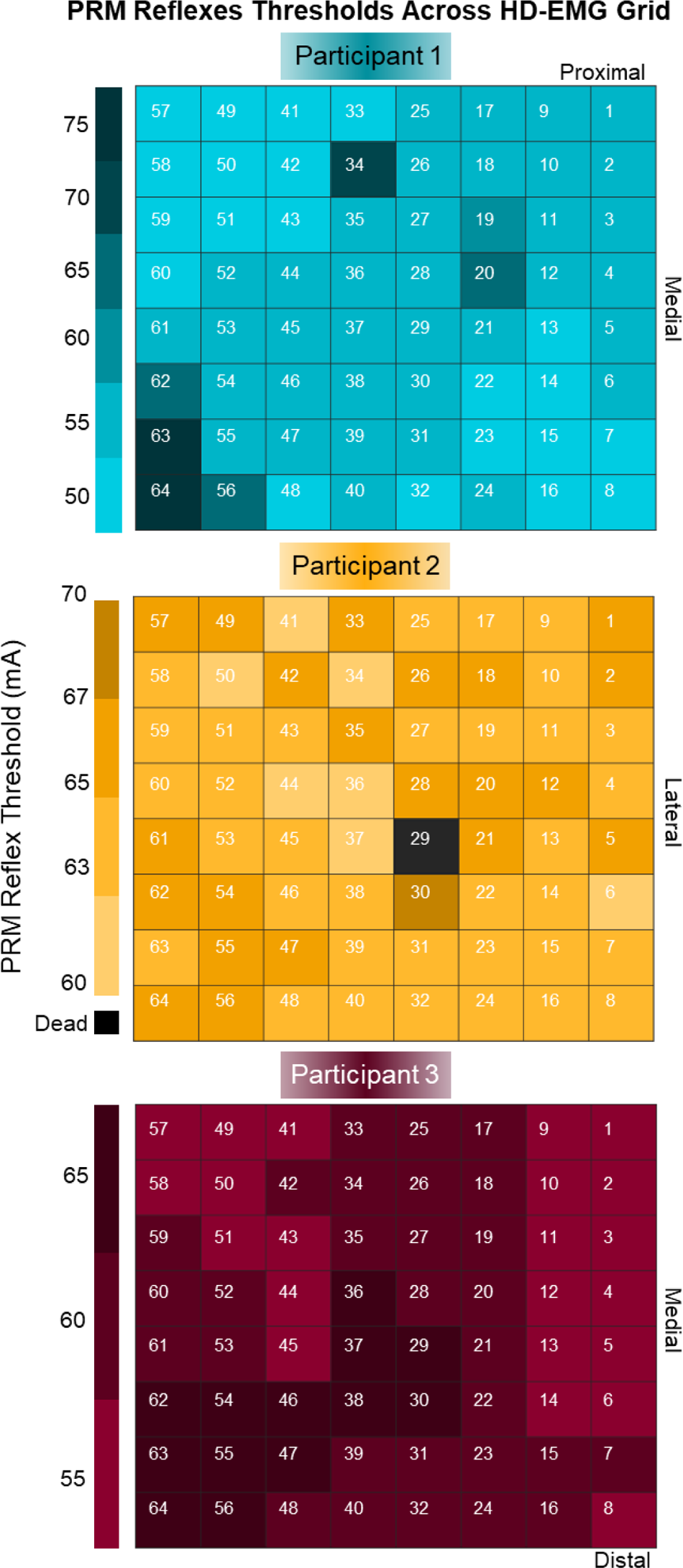
Threshold stimulation amplitudes for evoking a posterior root-muscle (PRM) reflex for each electrode on the high-density electromyography (HD-EMG) grid for each participant. The medial and lateral orientations of the grids are dependent on which leg was amputated.

**Supplementary Figure 3.**
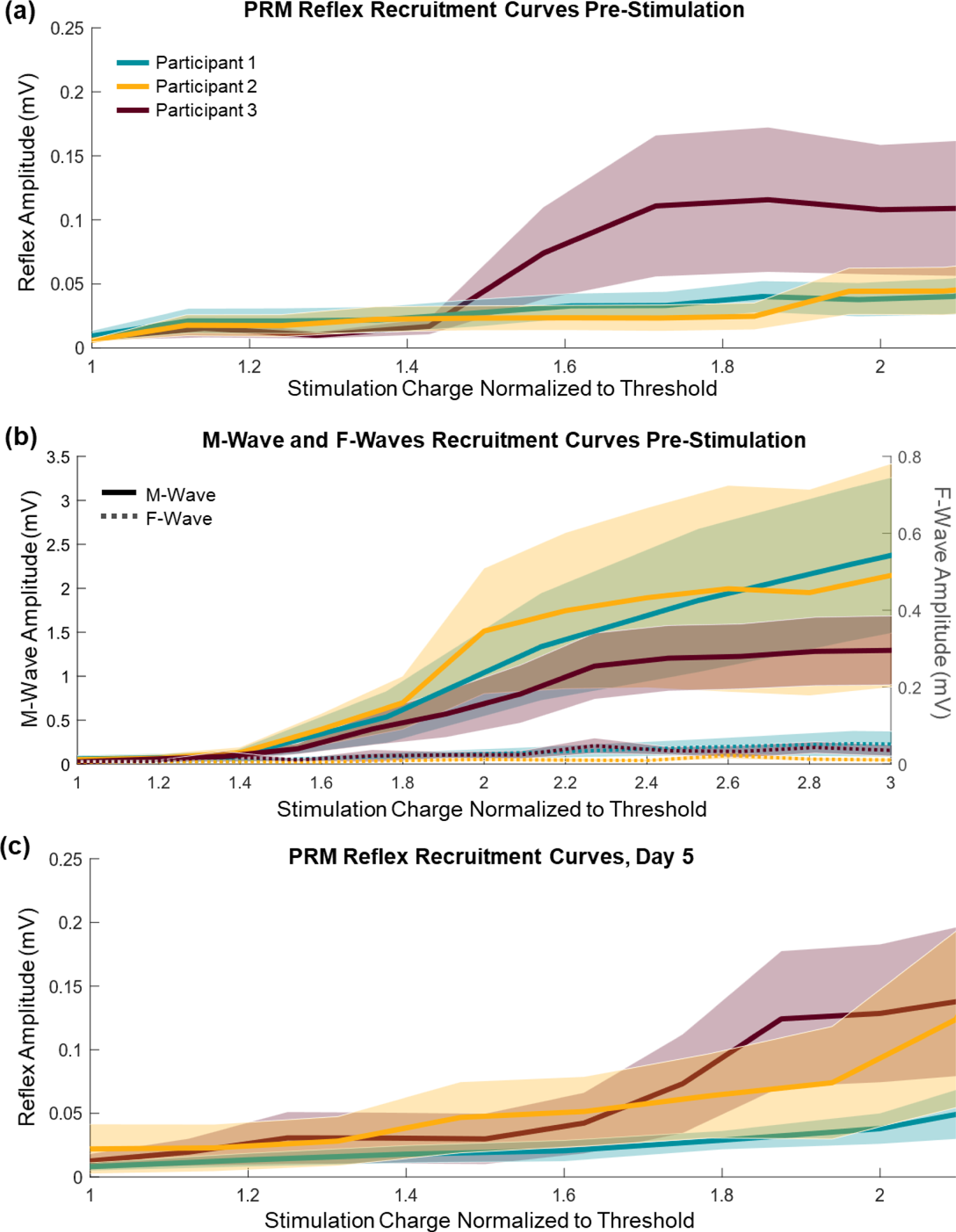
Recruitment curves, showing the peak-to-peak amplitude of the PRM reflexes as stimulation charge was increased, normalized to the threshold for evoking a PRM reflex for each participant, for (a) poster root-muscle (PRM) reflexes before stimulation, (b) M-waves and F-waves before stimulation, and (c) PRM reflexes after 5 days of stimulation.

**Supplementary Figure 4.**
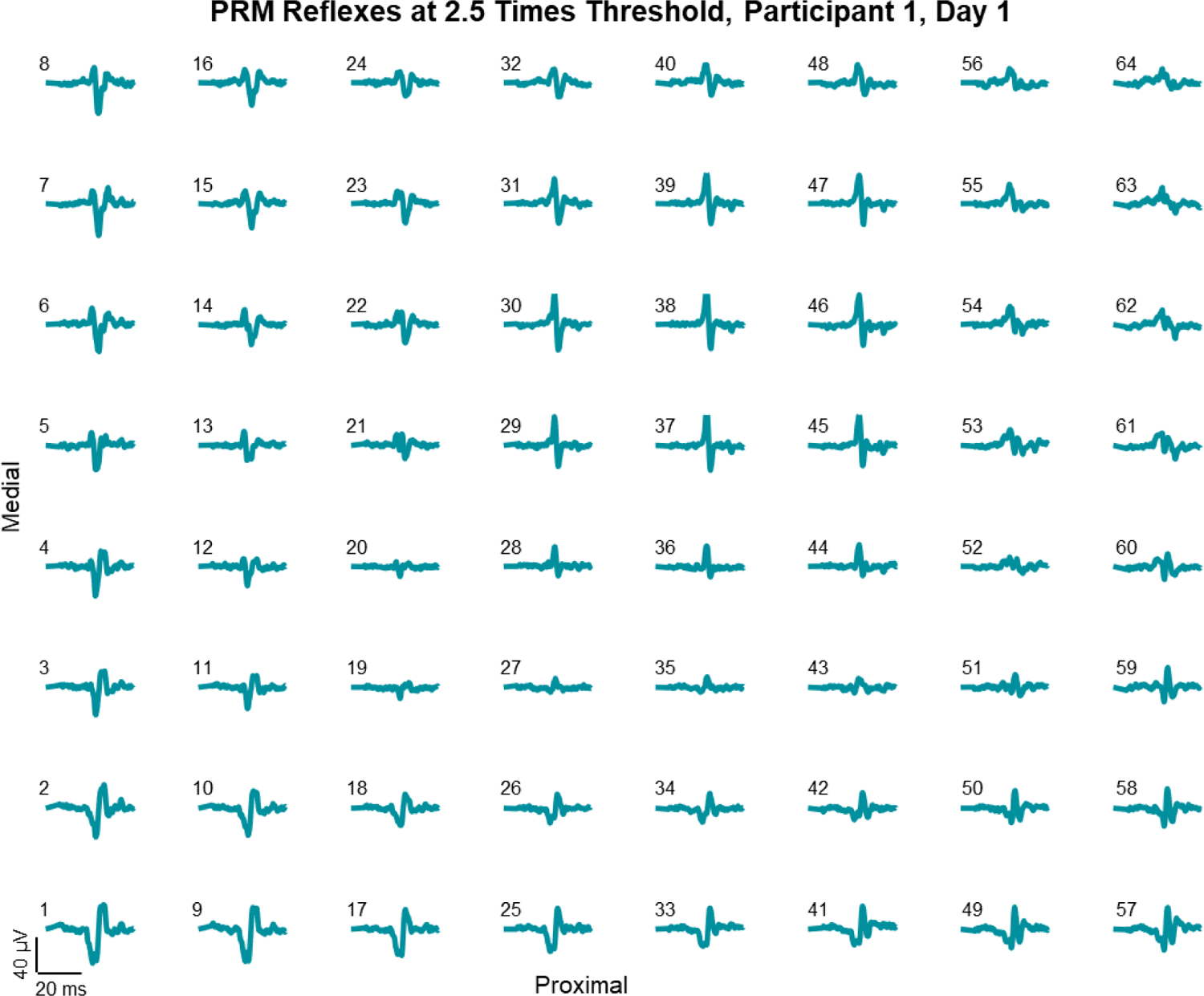
Examples of posterior root-muscle (PRM) reflexes recorded at each electrode on the high-density electromyography (HD-EMG) grid evoked at a stimulation amplitude 2.5 times higher than the mean threshold amplitude for Participant 1.

**Supplementary Figure 5.**
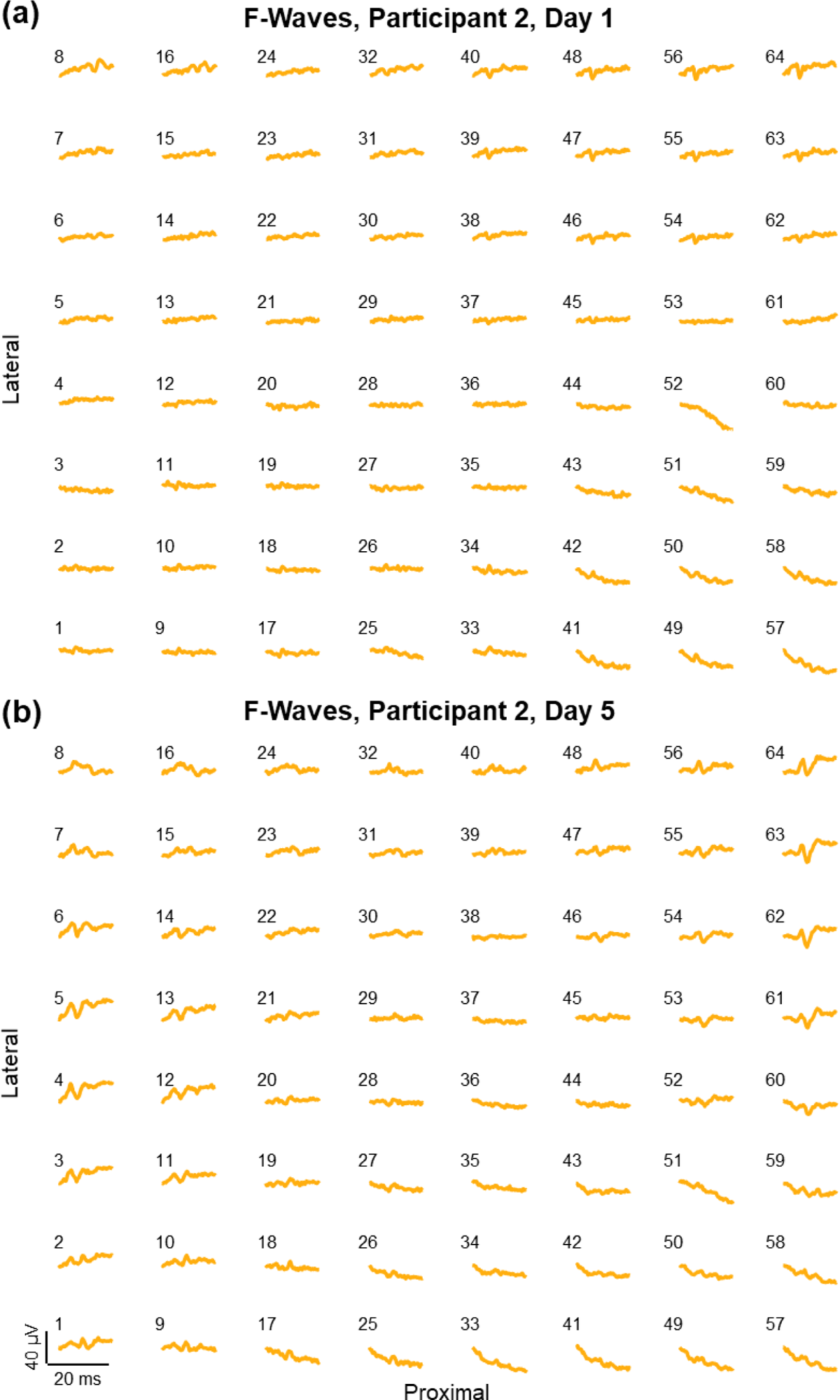
Examples of F-waves recorded at each electrode on the high-density electromyography (HD-EMG) grid evoked at maximal stimulation amplitude for Participant 2 on Day 1 (a) and Day 5 (b).

